# Willingness to volunteer of medical students during the COVID-19 pandemic: Assessment at a tertiary care hospital in India

**DOI:** 10.1101/2021.01.22.21250302

**Authors:** Manraj Sra, Amulya Gupta, Abhishek Jaiswal, Kapil Yadav, Anil Goswami, Kiran Goswami

## Abstract

**Background and Objectives:** The involvement of medical students in strategies to control COVID-19 might be considered to cope with the shortage of healthcare workers. This study aims at assessing the level of knowledge about COVID-19, willingness to volunteer, potential areas of involvement and reasons for deterrence towards volunteering among medical students.

**Methods:** A cross-sectional study was conducted among undergraduate medical students of a tertiary care teaching hospital in New Delhi. A web-based questionnaire was used to elicit demographic information, knowledge of COVID-19, willingness to volunteer and reasons for deterrence for working during COVID-19 pandemic and self-declared knowledge in six domains.

**Results:** A total of 292 students participated in the study with a mean age of 19.9±3.1 years. The mean (S.D.) knowledge score of COVID-19 was 6.9 (1.1) (maximum score 10). Knowledge score was significantly different among preclinical (6.5), paraclinical (7.18), and clinical groups (7.03). Almost three fourth (75.3%) participants were willing to volunteer in COVID-19 pandemic, though 67.8% had not received any training in emergency medicine or public health crisis management. Willingness to work was maximum in areas of social work and indirect patient care (62.3% each). Lack of personal protective equipment was cited as a highly deterrent factor for volunteering (62.7%) followed by fear of transmitting the infection to family (45.9%), fear of causing harm to the patient (34.2%), and absence of treatment (22.2%).

**Interpretation & conclusions:** Majority of the students were willing to volunteer even though they had not received adequate training. Students may serve as an auxiliary force during the pandemic, especially in the non-clinical setting.

## Introduction

COVID-19, a viral pneumonia caused by SARS-CoV2 has significantly overburdened healthcare facilities worldwide since its origin in December 2019. As compared to a country like India with just 43 healthcare workers (HCWs) per 10,000 population, the US and UK with 682 and 664 HCWs per 10,000 people respectively are much better equipped to handle the pandemic. [1] Despite this, healthcare systems worldwide have adopted various strategies like enrolling retired doctors, nurses and cancelling elective procedures to expand the medical surge capacity. [2] Diversion of HCWs to COVID-19 management has further affected routine care for other acute and chronic diseases. [3]

Medical students could potentially play a role in supporting HCWs during a pandemic, the precedent for which was during the 1918 Spanish Flu when medical students at the University of Pennsylvania helped in patient care. [4] Similarly, during a polio outbreak in Denmark in 1952, a group of students was tasked with manual ventilation of patients. [5] The United Kingdom and Italy have allowed final year medical students to graduate early and join the COVID-19 hospitals. Some schools in the US like the New York University followed suit. [6] Medical students at Harvard Medical School have made a “COVID-19 Student Response Team” to carry out tasks like supporting medical personnel in daily lives, spreading awareness and activism in the community. [7] In India, the government of Gujarat have instituted strategies to involve medical, dental, and nursing students in the COVID-19 response.[8]

With limited involvement in care delivery, students are seen as a non-essential part of clinical practice. However, medical students will become future healthcare workers and their involvement in COVID-19 care can be a valuable training experience. Further, being young adults, medical students are much less susceptible to COVID-19 related mortality as compared to retired healthcare workers, who are elderly with many having significant comorbidities. [9] This study aims at assessing the level of willingness to volunteer, knowledge about COVID-19, potential areas of involvement and reasons for deterrence to volunteering among medical students towards volunteering .

## Methods

A cross-sectional study was conducted using a web-based (Google Forms) questionnaire among all 475 undergraduate medical students of a tertiary care teaching hospital in New Delhi. The survey was sent out first on 24^th^ April 2020, two reminders were sent at a one-week interval. A semi-structured questionnaire was developed to analyze the medical student’s demographics, knowledge of COVID-19 pandemic, willingness to volunteer for working in COVID-19 pandemic, an inclination to work in various fields during COVID-19 pandemic (**a**. direct patient care, **b**. indirect patient care, **c**. laboratory work, **d**. social work, **e**. spreading awareness, **f**. supporting medical staff) and self-declared knowledge in those fields, and their reasons for deterrence. The questionnaire consisted of 37 questions.

There was information on the purpose of the study and question on consent at the beginning of the questionnaire, and the questionnaire will show only if the student gave the consent.

The face validity and content validity of the questionnaire were checked by one senior resident and 3 faculty members at the Department of Community Medicine of AIIMS, New Delhi. The questionnaire was pretested.

The correct responses for knowledge questions on COVID-19 were based on information provided by the Centre for Disease Control, World Health Organization, and ministry of health and family welfare (MoHFW). [10,11] Knowledge of COVID-19 was assessed using multiple-choice and short descriptive questions. A 6-point Likert scale was used to assess the inclination to work and their self-declared knowledge in 6 areas namely: history taking through phone, direct patient care, spreading awareness, supporting health care workers outside the hospital, laboratory work, and social work. Deterrence factors for volunteering were assessed using matrix-based question with 6 potential reasons for deterrence to be graded as highly deterrent, moderately deterrent, weakly deterrent, and not deterrent. The respondents were asked the maximum mortality rate ranging from 0% to 100% at which the students were willing to work under two scenarios - with minimal safety equipment (MS) or with full-body safety equipment (FS). Two open-ended free text questions were asked for any reasons for deterrence and potential areas of involvement that were not listed in the survey.

The students were classified into pre-clinical, para-clinical and clinical years depending on their current year in the course. The pre-clinical year consisted of MBBS students who were currently in 1st year, para-clinical year of MBBS students who were currently in the 2nd-year and clinical year of MBBS students who are currently in 3rd year, 4th year, or were interns.

For knowledge assessment, a score out of 10 was given to each respondent depending on the number of correct answers. Responses involving the Likert scale (1-6) for self-declared knowledge and inclination to work was dichotomized: 1-3 indicating Not enough knowledge/ Not willing to volunteer and 4-6 indicating Enough knowledge/Willing to volunteer. A multivariate logistic regression model was developed to capture the relationship between the willingness to work and (i) gender, (ii) year of study, and (iii) knowledge of COVID-19. All variables with p-value less than 0.25 were entered in the multivariate model. Odds ratio were provided with 95% confidence intervals.

The model included all subjects with complete data on all variables. Since the responses to all question that were included in the model were compulsory no response was excluded due to incomplete data. All p-values are two-tailed and a p-value of less than 0.05 was considered to be significant. Statistical analysis was performed using STATA version 13.1 (StataCorp LP, College Station, TX, USA).

The study was approved by the Institute Ethics Committee (ID-IEC/267/17.04.2020).

## Results

Two hundred ninety-two participants gave consent and responded. The overall response rate was 61.5%, comprising 292 respondents, including 85 from the pre-clinical year (79.4% response rate), 90 from para-clinical years (84.1% response rate), 107 from clinical years (40.9% response rate). 76.4% of the respondents were male with a mean age of 19.9 ± 3.1 years. The demographic details and previous training and awareness of the students have been summarized in Table 1.

**Table 1:** Demographic details and previous training of our study population.

|  |  |  |
| --- | --- | --- |
| **Gender % (N)** | Male | 76.37 (223) |
|  | Female | 23.63 (69) |
| **Mean age (in years)**<br>**(Mean ± SD)** | | 19.9± 3.1 |
| **Study Year,**<br>**% (N)** | Pre-clinical years (*1st year*) | 29.11 (85) |
|  | Para-clinical years (*2nd year*) | 30.82 (90) |
|  | Clinical years (*3rd year, 4th year and Interns*) | 40.07 (107) |
| **Previous training,**<br>**% (N)** | Basic life support | 30.5 (89) |
|  | Triage | 4.8 (14) |
|  | Infection control measures | 8.9 (26) |
|  | Handling hazardous material like Bio-medical waste in hospital | 13.4 (39) |
|  | Management of suspected COVID-19 cases | 2.4 (7) |
|  | Public health emergency | 1.4 (4) |
|  | Other | 2.4 (7) |
|  | Not received any of the above training | 67.8 (198) |

### Knowledge and Skills

The overall mean (S.D.) knowledge score of COVID-19 among participants was 6.92 (1.05) (out of a maximum score of 10). Distribution of mean knowledge score according to study year groups was as follows: pre-clinical years had a mean (S.D) knowledge score 6.5 (1.12), paraclinical and clinical years had a score of 7.18 (0.97) and 7.03 (0.99) respectively. The knowledge score was significantly different among groups (One-way ANOVA, F-ratio = 10.67, p <0.001). Most of the participants (98.63 %, 288) were aware of COVID-19 pandemic situation. Almost half of the respondents (48.29%, 141) reported being aware of the clinical management of a COVID-19 patient, 59.59% (174) reported knowing appropriate use of personal protective equipment (PPE), and 70.55% (206) were aware of mitigation procedures like contact tracing, etc. 67.8% of the respondents had not received any training in emergency medicine or public health crisis management. More than three-fourth (75.3%) participants indicated their willingness to volunteer in the COVID-19 pandemic.

### Knowledge and Inclination to Work

The inclination to work and self-declared knowledge of 6 domains was analyzed, they were as follows: direct patient care included assisting in minor procedures or screening of patients; indirect patient care involved telemedicine; laboratory work included assisting in COVID-19 testing laboratories; social work included helping out with governmental and non-governmental agencies to cater to the underprivileged; spreading awareness in the community through social media or other means, and supporting medical staff in their households.

The average inclination to work was highest for social work and indirect clinical work (62.3% for both) followed by lab work (51.7%), spreading awareness (48.3%), direct clinical work (47.6%), and supporting staff non-medically (39.4%). The self-declared knowledge level was highest for supporting staff non-medically (55.48%) followed by spreading awareness (54.11%), indirect clinical work (52.05%), social work (43.49%), laboratory work (33.22%) and direct clinical work (24.69%). All the domains showed a positive and significant correlation between knowledge of the participant and inclination to work (p <0.01). The inclination to work and self-declared knowledge for each domain have been summarized year wise in Figure 1a and Figure 1b respectively.

**Figure 1.**
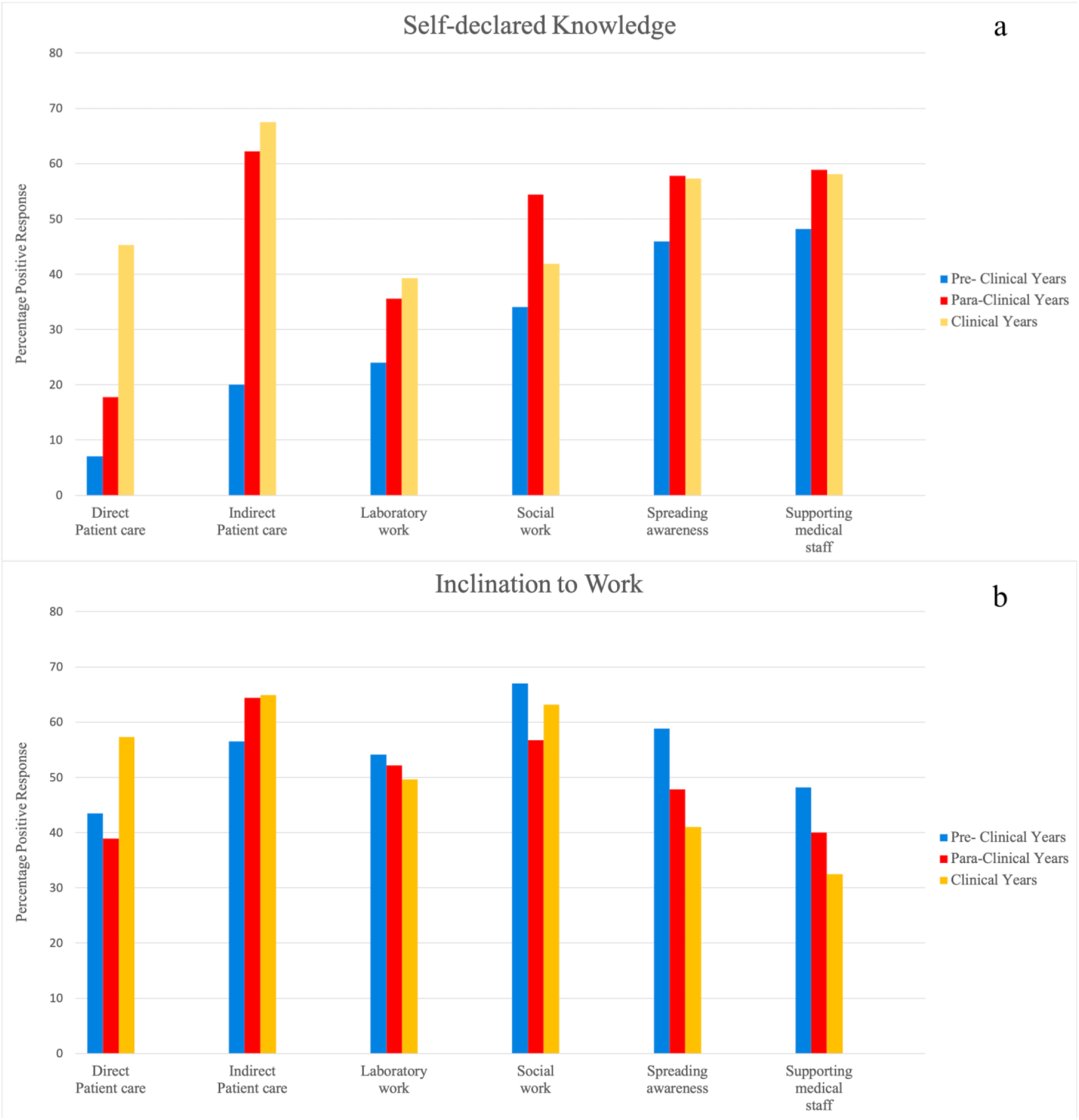
a: Proportion of medical students who showed positive inclination to volunteer in different domains. b: Proportion of medical students who showed sufficient knowledge in different domains.

### Reasons for deterrence

The lack of PPEs was seen to be the most deterrent factor (rated highly deterrent by 67.2% of respondents), the fear of watching someone die was rated as the least deterrent factor (rated highly deterrent by 5.5% of respondents). Preclinical students showed greater deterrence due to parental factor (p=0.033) and fear of watching someone die (p=0.026) as compared to students of para-clinical and clinical years. Responses for all reasons have been summarized in Figure 2. The respondents were asked the maximum mortality rate of a disease ranging from 0% to 100%, at which they were willing to volunteer under two situations, one where minimal safety equipment (MS) was provided and second where full safety equipment including bio-hazard suits (FS) was provided (Table 2). These values signify the fear for one’s safety during volunteering. The students in the pre-clinical, para-clinical and clinical years indicated a median threshold of 5%, 2 % and 1 % respectively in MS conditions. Comparatively, in the FS conditions, the students of pre-clinical, para-clinical and clinical years indicated a median threshold of 10 %, 8 % and 5 % respectively. There was a significant difference between the maximum mortality rates of MS and FS (p < 0.01). 37 students filled the open-ended question for the reasons for deterrence. Incidents of violence against health professionals were the most prominent theme. This was followed by apathy or lack of concern for the well-being of others.

**Table 2:** Willingness to volunteer with respect to mortality and study year.

| Maximum mortality of a communicable disease at which you may volunteer | Pre-Clinical<br>Median (IQR) | Para-Clinical<br>Median<br>(IQR) | Clinical<br>Median<br>(IQR) |
| --- | --- | --- | --- |
| If minimal safety equipment is provided | 5 (1-25) | 2 (1-5) | 1 (0.02-5) |
| If full safety equipment is provided | 10 (2-50) | 8 (3-16) | 5 (1-16) |

**Figure 2:**
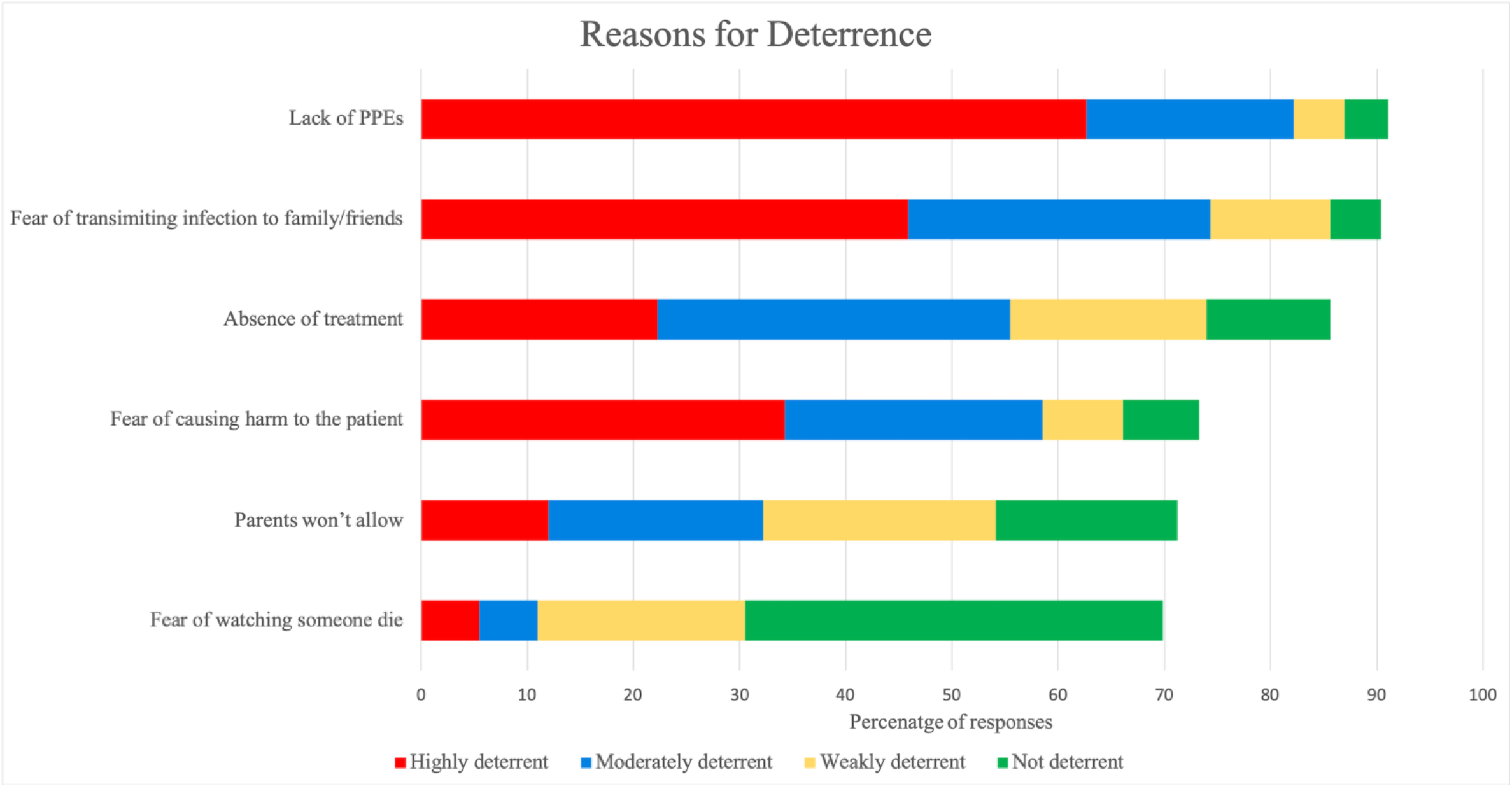
Reasons for deterrence to volunteer in COVID-19 pandemic situation.

The open-ended question for additional ways to contribute to the pandemic was filled by 36 students. Contact tracing was the only new theme amongst the answers given.

In comparison to students of pre-clinical years, the students of para-clinical (aOR=1.61 (0.79-3.29), p= 0.193) and clinical years(aOR=1.23 (0.64 - 2.39), p=0.538) were more likely to volunteer but the difference was not significant (Table 3) Female respondents (aOR=2.09 (0.98 - 4.47), p=0.057) were also found to be more willing to volunteer as compared to male respondents. The knowledge of COVID-19 did not significantly affect the willingness to volunteer of the respondents (aOR=1.12(0.86-1.46), p=0.396).

**Table 3:** Multivariate logistic regression model for willingness to volunteer.

| Variable |  | Unadjusted OR<br>(95% C.I.) | P-value | Adjusted OR<br>(95% C.I.) | P-value |
| --- | --- | --- | --- | --- | --- |
| Year | Pre-clinical year | Ref | - | Ref | - |
|  | Para-clinical year | 1.86 (0.93-3.71) | 0.077 | 1.61 (0.79 - 3.29) | 0.193 |
|  | Clinical Year | 1.55 (0.83- 2.91) | 0.170 | 1.23 (0.64 - 2.39) | 0.538 |
| Gender | Male | Ref | - | Ref | - |
|  | Female | 2.27 (1.09-4.72) | 0.028 | 2.09 (0.98 - 4.47) | 0.057 |
| Knowledge score (continuous scale) |  | 1.22 (0.95-1.58) | 0.124 | 1.12 (0.86 - 1.46) | 0.396 |

## Discussion

Medical students can be a potentially important resource in this public health crisis. Previously limited work has been done to access the capabilities, attitudes, and concerns of medical students towards volunteering in such a situation.

The present study found that majority of medical students across various years were willing to volunteer in pandemic COVID-19 situation. They were most inclined to work in areas of indirect patient care and social work. The most important reasons for deterrence to volunteer was the lack of PPE and the fear of transmitting the disease to family and friends. Another key observation is that a considerable number of medical students were aware of various aspects of management of a pandemic though they have not received any formal training to deal with a public health emergency.

On stratification of students according to their year of study, key observations are brought out in their attitudes towards volunteering. Students in the pre-clinical years indicated greatest inclination to work in social work and spreading awareness. This is in the agreement with the fact that with the limited clinical knowledge, they are best suited for non-clinical work. Students of the para-clinical years were most inclined to work in indirect patient care and social work. Students of clinical years showed the highest inclination to work in direct and indirect patient care. These observations correlate with the level of knowledge of each group. Thus, these trends show us that if the need arises for medical students to be involved in the management of the pandemic stratification of students is important for effective utilization of their services.

The leading reasons why students were hesitant to volunteer were lack of PPEs, fear of spreading the disease to family and friends and absence of treatment. These concerns agree with previous studies. [12,13] This is also supported by the fact that students reported a significantly higher mortality rate at which they were willing to work when provided with full safety gear as compared to conditions with minimal safety gear. These concerns might be exacerbated by the fact that medical students think of COVID-19 as one of the deadliest coronaviruses, as indicated in a study at Karachi. [14] Since our study was conducted during the initial months of the outbreak the fear of lack of PPE was significant, but with progress of the outbreak most place have had sufficient availability of PPE. Another factor that deters students is the fear of causing harm to the patient, this should be taken into consideration especially when involving students in direct patient care. Previous studies have also shown that medical students have poor knowledge of patient safety. [15] Studies have also shown that in the event of students causing any harm, they might endure considerable stress and even quit work. [16] Deterrence towards volunteering due to familial reasons was seen more in students of pre-clinical years this factor is considerably dependent on the societal perception of threat from COVID-19. Fear of watching someone die, though a minor factor for deterrence should in no way be ignored as previously it has been shown that students who have worked in emergency medicine may suffer from negative psychological effects of such exposure. [17]

Previously, studies conducted at the University of Alberta before the influenza H1N1 showed that more than half of the students believed that medical students have an obligation to be involved in influenza pandemics. [18]Another study conducted after the H1N1 pandemic at the University of Michigan showed that 88% of the students preferred to be formally involved in the response to this crisis. [19]

Willingness to volunteer was significantly higher in females (χ2 = 5.025, p = 0.025) as compared to males. This supports the findings of previous studies done among hospital volunteers [20] and medical students [21] which found female HCWs were more dependable at the times of crisis. However, this was in contradiction to the results of a study done at the University of Alberta [22] which showed no significant impact of gender on willingness to volunteer in Canadian students. The maximum mortality of a communicable disease at which the student is willing to volunteer showed a decreasing trend from preclinical to clinical years. This reflects the fact that without moral training, idealism in medical students tends to decrease over the years. [23]

In contrast to a study at Rutgers University [24] where a mortality threshold of 34% for respiratory illnesses was found, our values were visibly lower. This may be due to a difference in survey questions. (Present study questionnaire prompted the participant to write any value while that of Rutgers university used increments of 10% as options), exacerbated by fear during the ongoing pandemic or demographic differences (lesser female respondents and only students of the Indian subcontinent in our study).

In understanding the implications of this study, a few limitations must also be considered. Our study was a single-centre study, different institutes may have a varying emphasis on emergency medicine and public health crisis management. Moreover, filling all options in the reasons for deterrence section was not compulsory. Limitations in data analysis include dichotomizing the ordinal questions and depending on self-reported information could bias the results as well. Due to a lower response rate for the students in the clinical years, the findings might not be generalizable. However, it is known that the response rate is generally low in clinical healthcare workers [25] which does not change significantly with the mode of administration. [26]

Despite the limitations, our study provides considerable insights about the knowledge, skills and attitudes of medical students in a public health crisis. The ethical and moral issues surrounding the involvement of medical students in such a pandemic require deeper probing. Our study helps in identifying the key areas where medical students would be most comfortable in being involved and their reasons for deterrence to volunteer. Ultimately, the findings highlight that medical students are an untapped resource that may serve as a valuable resource despite their limited skill set, and the critical role of disaster management programs in medical schools that will serve to translate the inclination of volunteering of students to actual service.

## Data Availability

Anonymized responses of the participants will be shared on a reasonable request to the corresponding author.

## Authors contribution

MS and AG were involved in the designing of the study, data collection and writing the manuscript. AJ was involved in data analysis and writing the manuscript. KY, AG and KG supervised the study.

